# Evaluation of Photon-Counting CT for Spectral Imaging in Cardiovascular Applications: Impact of Lumen Size, Dose, and Patient Habitus

**DOI:** 10.1101/2025.01.07.25320150

**Authors:** Martin V. Rybertt, Leening P. Liu, Manoj Mathew, Pooyan Sahbaee, Harold I. Litt, Peter B. Noël

## Abstract

**Objectives:** This study evaluates the performance of a clinical dual-source photon-counting computed tomography (PCCT) system in quantifying iodine within calcified vessels, using 3D- printed phantoms with vascular-like structures lined with calcium.

**Methods:** Parameters assessed include lumen diameters (4, 6, 8, 10, and 12 mm), phantom sizes (S: 20×20 cm, M: 25×25 cm, L: 30×40 cm, XL: 40×50 cm, representing the 99th percentile of US patient sizes), and iodine concentrations (2, 5, and 10 mg/mL). Scans were performed at radiation dose levels of 5, 10, 15, and 20 mGy to systematically evaluate iodine quantification accuracy and spectral imaging performance.

**Results:** The results indicate that for lumen diameters ≥6 mm, iodine quantification remains stable across all dose levels and phantom sizes, with deviations consistently below 0.6 mg/mL. Whereas, for 4 mm lumens, stability is observed primarily in smaller to medium phantoms, highlighting the influence of patient size and radiation dose on quantification accuracy. Virtual Monoenergetic Imaging (VMI) at 70 keV showed stable performance for larger lumens (≥6 mm) with variations of 13 ± 2 HU across all conditions, while smaller lumens remained stable in medium to small phantoms.

**Conclusions:** These findings highlight the influence of lumen diameter, patient size, and radiation dose in optimizing PCCT protocols for spectral imaging. Importantly, the study demonstrates that PCCT delivers stable and highly accurate imaging across nearly the entire range of patient sizes in the U.S..

**Advances in knowledge:** This study demonstrates PCCT’s potential to enhance spectral imaging in vascular applications, surpassing conventional or Dual Energy CT.

## Introduction (450)

Coronary Artery Disease (CAD) is the leading cause of death in the United States, with over 300,000 fatalities reported in 2023^1^. This high mortality rate underscores the critical need for effective diagnostic tools and early detection methods. Currently, one of the primary diagnostic tools for CAD is computed tomography (CT), which has been associated with a reduction in cardiovascular mortality^2^. Diagnosing CAD presents significant challenges due to the small diameters of coronary vessels and the difficulty in differentiating between calcified plaque and iodinated contrast^3–5^. Traditional CT imaging may not provide the necessary contrast resolution to distinguish these accurately, leading to potential diagnostic ambiguities. Spectral CT addresses this issue by leveraging the attenuation characteristics of different materials to produce material-specific reconstructions^6,7^. For example, Spectral CT can generate maps that isolate iodine, thereby enhancing the visibility of contrast agents used during imaging. However, despite these advances, there are still challenges to overcome and differentiating between materials with similar attenuation values, such as calcium and iodine, remains a significant obstacle^8^.

The clinically available dual-source PCCT represents an advance over other Spectral CT technology by offering improved material decomposition, reduced radiation dose, and enhanced spatial resolution, making it particularly well-suited for cardiovascular applications^9–11^. For diagnosing CAD, this scanner uniquely combines these improvements with the temporal benefits of dual-source imaging^12^. Studies have already explored PCCT’s applications for CAD diagnosis showing that it provides greater diagnostic accuracy than other types of Spectral CT^13^, especially in larger patients, where it offers improved detectability compared to conventional CT^14^. PCCT has also been used to assess stent and plaque composition^15,16^, demonstrating an ability to reduce blooming artifacts and providing a clearer view of stent structures^17,18^. Additionally, PCCT has been evaluated for calcium scoring directly from contrast enhanced scans, thus reducing radiation exposure while achieving performance comparable to noncontrast CT^19–22^. While PCCT has demonstrated strong diagnostic capabilities, with multiple studies highlighting its advantages in technical performance evaluations^23,24^, a critical question for CAD diagnostics remains: can PCCT accurately quantify iodine in relation to vessel diameter and calcium presence. The spatial resolution in the spectral mode of the clinically approved PCCT provides high-quality spectral data at a resolution superior to that of conventional CT. Although the absolute quantification of iodine may be secondary in CAD diagnostics, its reliable association with virtual non-contrast imaging is crucial for detecting occlusions. The performance of virtual non-contrast imaging directly depends on accurate iodine quantification, as iodine removal requires precise determination of its concentration.

In this study, we aim to evaluate the complex relationships among target / lumen size, patient habitus, iodine concentration, and radiation dose in influencing the accuracy of spectral results with a focus on iodine quantification. By systematically analyzing how these factors interact, we seek to better understand their combined impact on iodine measurement reliability, particularly in relation to CAD diagnostic accuracy.

## Methods (603)

### Phantom Design

A set of 3-D printed phantoms were designed for use as inserts into a commercially available technical phantom (Multi-energy CT Phantom, Sun Nuclear, Melbourne, FL, USA) to evaluate the performance of iodine quantification in small calcified coronary vessels. All phantoms were printed (TAZ Sidekick, Lulzbot, Fargo, ND, USA) using a calcium-doped polylactic acid (PLA) filament (StoneFil, FormFutura, AM Nijmegen, The Netherlands) to mimic coronary calcifications. Each 3-D printed insert contains a constant diameter u-shaped tube that can be filed with iodinated contrast. Moreover, each tube has a lumen diameter ranging from 4 to 12 mm (4, 6, 8, 10, 12 mm) and includes a calcium lining of 1 mm with a Hounsfield unit of 380±24 HU to imitate the anatomy of coronary vessels with advanced atherosclerosis.

### Image Acquisition

The 3-D printed phantoms were scanned on a first-generation clinical dual-source PCCT (NAEOTOM Alpha, Siemens Healthineers, Munich, Germany) within the technical phantom consisting of various tissue-mimicking and material-specific inserts and an outer extension ring. To determine how patient habitus impacts iodine quantification, the 3-D printed inserts were scanned in different sized phantoms. The smallest setup (S) used the inner part of the technical phantom with a diameter of 20 cm. For the medium (M) setting, the S phantom was positioned within a 3-D printed PLA ring, increasing the diameter to 25 cm. The large (L) setting was created by placing the inner phantom inside the 30×40 cm extension ring of the technical phantom. For the extra-large (XL) setting, the L phantom was placed in a 3-D printed 40×50 cm outer ring, as illustrated in Figure 1. Note that the XL setting is equivalent to the abdominal size of the 99th percentile of US patients.

**Figure 1.**
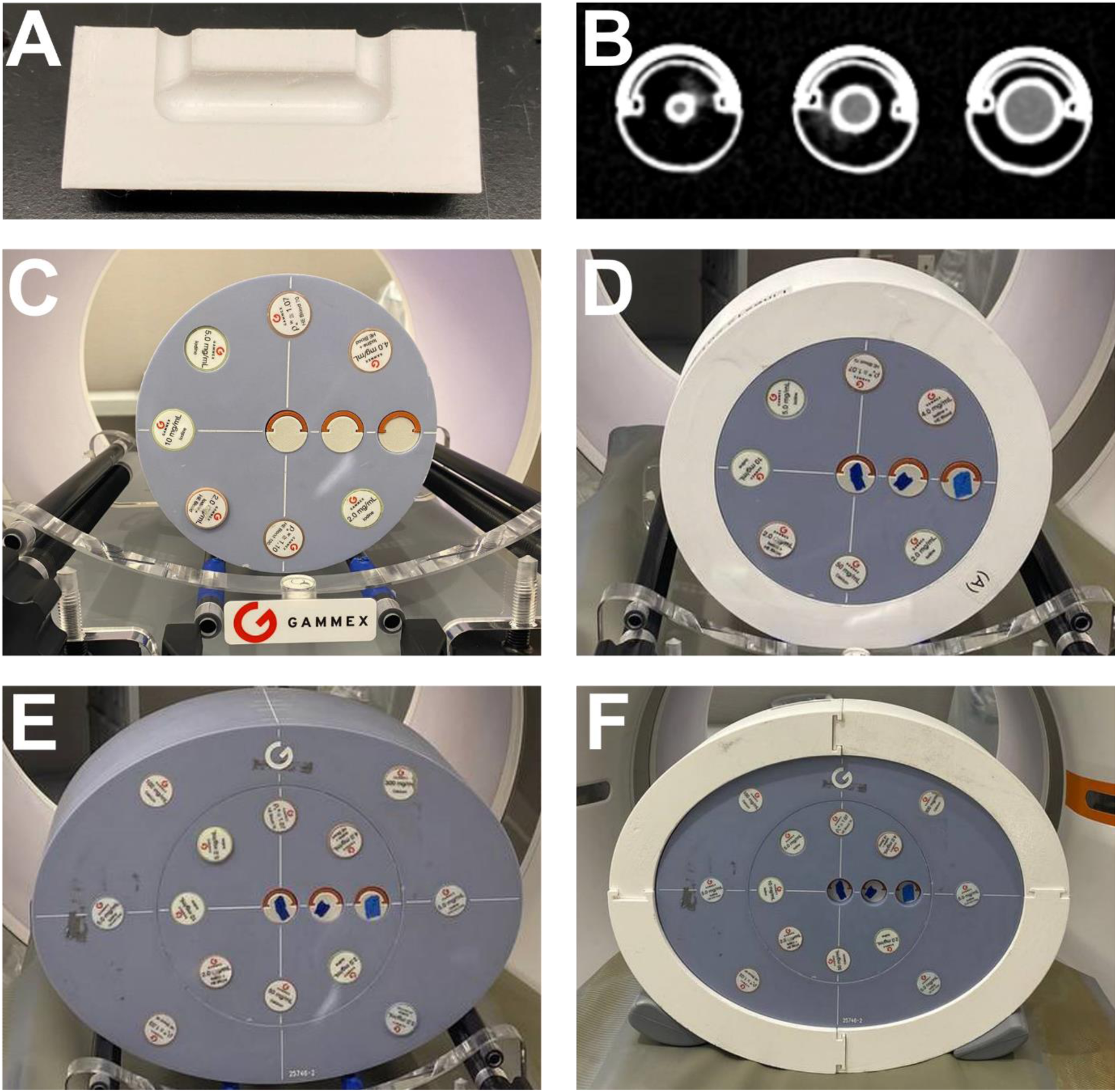
Experimental setup on PCCT scanner. Phantom rod inserts consist of a constant diameter u-shaped tube that can be filled with contrast material (A) with each insert having a different diameter and a 1 mm calcium lining (B). Rods were scanned in four separate simulated patient sizes: small (C), medium (D), large (E), and extra-large phantoms (F).

After filling the inserts with a 2 mg/mL iodine solution (Isovue-300, Bracco Diagnostics, Milan, Italy) and positioning them within each phantom configuration, scans were conducted at a tube voltage of 120 kVp across four volumetric CT dose index (CTDI_vol_) levels (5, 10, 15, and 20 mGy). Each dose level was repeated three times to evaluate reproducibility (Table 1). Subsequently, this procedure was repeated for higher iodine concentrations (5 mg/mL and 10 mg/mL) to assess the effects of varying contrast levels on quantification accuracy.

**Table 1.** Acquisition and reconstruction parameters.

| Parameter |  |
| --- | --- |
| Iodine concentration [mg/mL] | 2, 5, 10 |
| CTDI<sub>vol</sub> [mGy] | 5, 10, 15, 20 |
| Phantom size [cm] | 20x20, 25x25, 30x40, 40x50 |
| Spectral results | VMI 70 keV, ID, VNC |
| Tube Voltage [kVp] | 120 |
| Pitch | 1 |
| Acquisition Mode | Helical, Non-gated |
| Reconstruction Kernel | Qr40 |
| Slice thickness [mm] | 3 |
| Field of View [mm] | 250 |

### Analysis

Spectral results were reconstructed for each scan, including virtual monoenergetic images (VMI) at 70 keV, iodine density (ID), and virtual non contrast (VNC). Reconstruction was performed with a field of view of 250 mm, a slice thickness of 3 mm, reconstruction filter of Qr40, and quantum iterative reconstruction level of 0. Regions of interest (ROI) of 60% of tube diameter were drawn manually for each insert using the VMI 70 keV images from the 20 mGy scans for each phantom size. ROI placement information was saved and utilized for the measurements of other spectral results for the same phantom size. The mean of each ROI was determined, and the corresponding variation in the mean was calculated as the standard deviation of the means across eight consecutive slices of three scans (24 slices total).

To illustrate the effects of phantom size and lumen diameter on image quality, VMI 70 keV slices were visually compared in the coronal plane. Scatter plots were generated to visualize the influence of lesion size, phantom size, iodine concentration, and dose for iodine density (ID), VMI, and VNC images with each point representing mean measurements from ROIs and their corresponding standard deviations across slices from all repetitions. Finally, to quantify deviations in lumen diameter, we calculated the absolute difference from a reference value, defined as the diameter of the 12 mm insert within the small phantom at a dose of 20 mGy.

## Results (929)

Spectral results in the coronal plane were used to assess the influence of phantom size and lumen diameter on image quality, with findings presented in **Figure 2**. These images illustrate a clear trend where variations in both phantom size and lumen diameter significantly affect visual clarity and structural detail resolution. Specifically, larger phantom sizes and smaller lumen diameters were associated with a reduction in image contrast, whereas smaller phantom sizes yielded improved delineation of the lumen boundaries.

**Figure 2.**
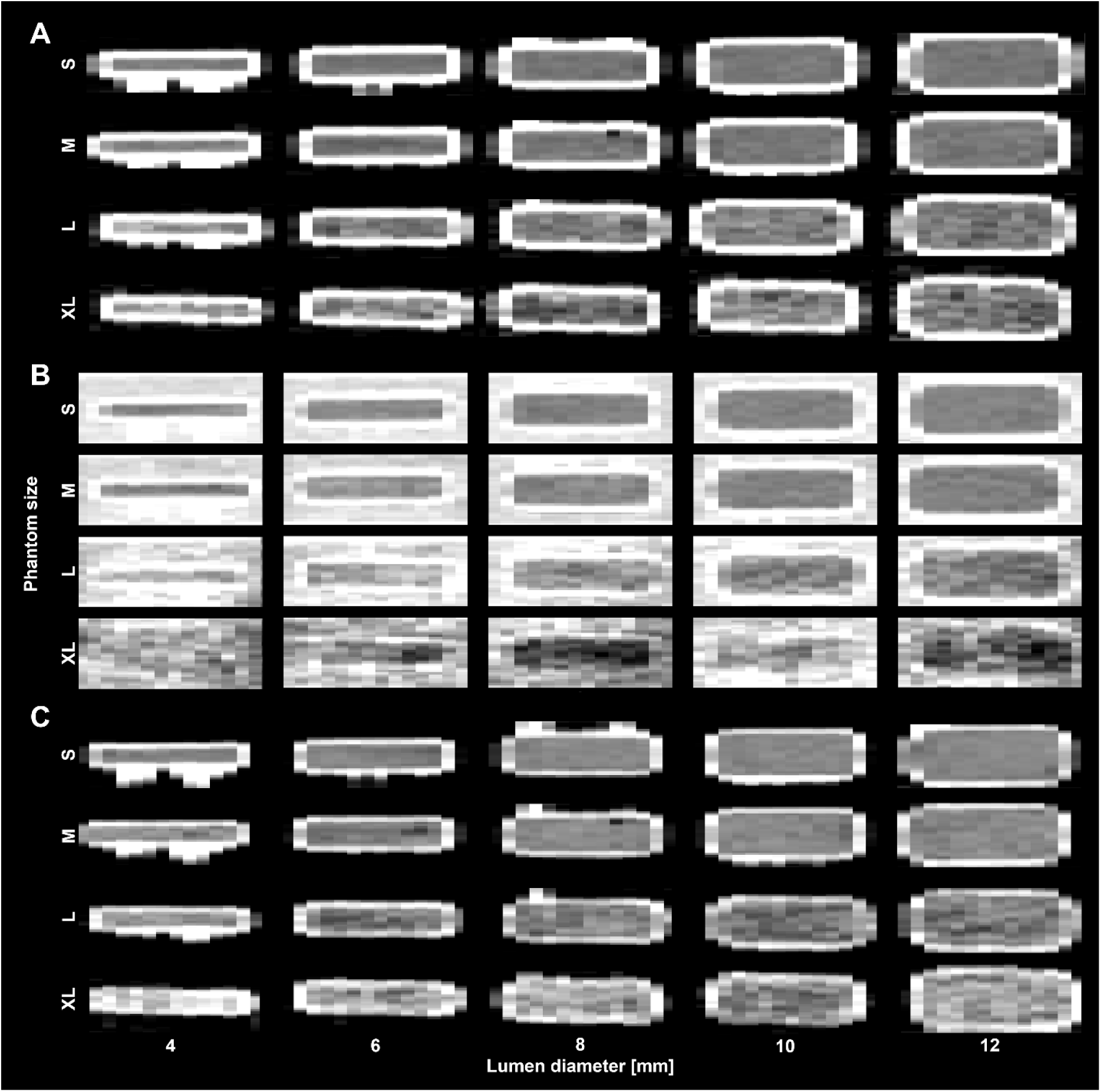
Coronal images of spectral results with varying phantom sizes and lumen diameter. 3-D printed phantoms were filled with a solution with an iodine concentration of 5 mg/mL at a radiation dose of 20 mGy. VMI 70 keV (A, WL/WW: 150/300 HU) exhibited consistent quantification with S, M, and L phantoms for lumen diameters greater than 6 mm. Contrast in iodine density images (B, WL/WW: 5/10 mg/mL) is easily discernible for S, M, and L phantom sizes with less conspicuous calcium shell than VMI. VNC images (C, WL/WW: 0/300 HU) have comparable quality to VMI, with increased noise at larger phantom sizes. Image quality decreased as phantom size increased.

The iodine density (ID) maps demonstrated good agreement with reference values for lumen diameters above 6 mm, again showing dependency on patient size, as illustrated in **Figure 3**. Specifically, ID maps exhibited high accuracy for lumen diameters greater than 6 mm at a dose level of 20 mGy. This improved in smaller phantoms (S and M), where accurate ID quantification was observed consistently across all tested iodine concentrations. For the S and M phantom sizes, the absolute relative differences in ID values remained minimal; the peak relative difference observed was 0.9 mg/mL. For the M phantom, the average differences were 0.1 ± 0.1 mg/mL, 0.3 ± 0.2 mg/mL, and 0.4 ± 0.3 mg/mL for iodine concentrations of 2 mg/mL, 5 mg/mL, and 10 mg/mL, respectively. These results indicate that in smaller phantoms, the ID maps achieve reliable accuracy even at low iodine concentrations. ID map accuracy demonstrated similar trends across all iodine concentrations in the larger L and XL phantom sizes; however, the larger sizes generally resulted in increased relative differences, particularly in the XL phantom. For instance, at an iodine concentration of 5 mg/mL, average ID differences were 0.5 ± 0.4 mg/mL and 1.3 ± 0.4 mg/mL for the L and XL phantoms, respectively. The XL phantom displayed significantly higher relative differences, suggesting, decreased precision of ID measurements in larger patient or phantom sizes, potentially due to increased scatter and decreased contrast resolution.

**Figure 3.**
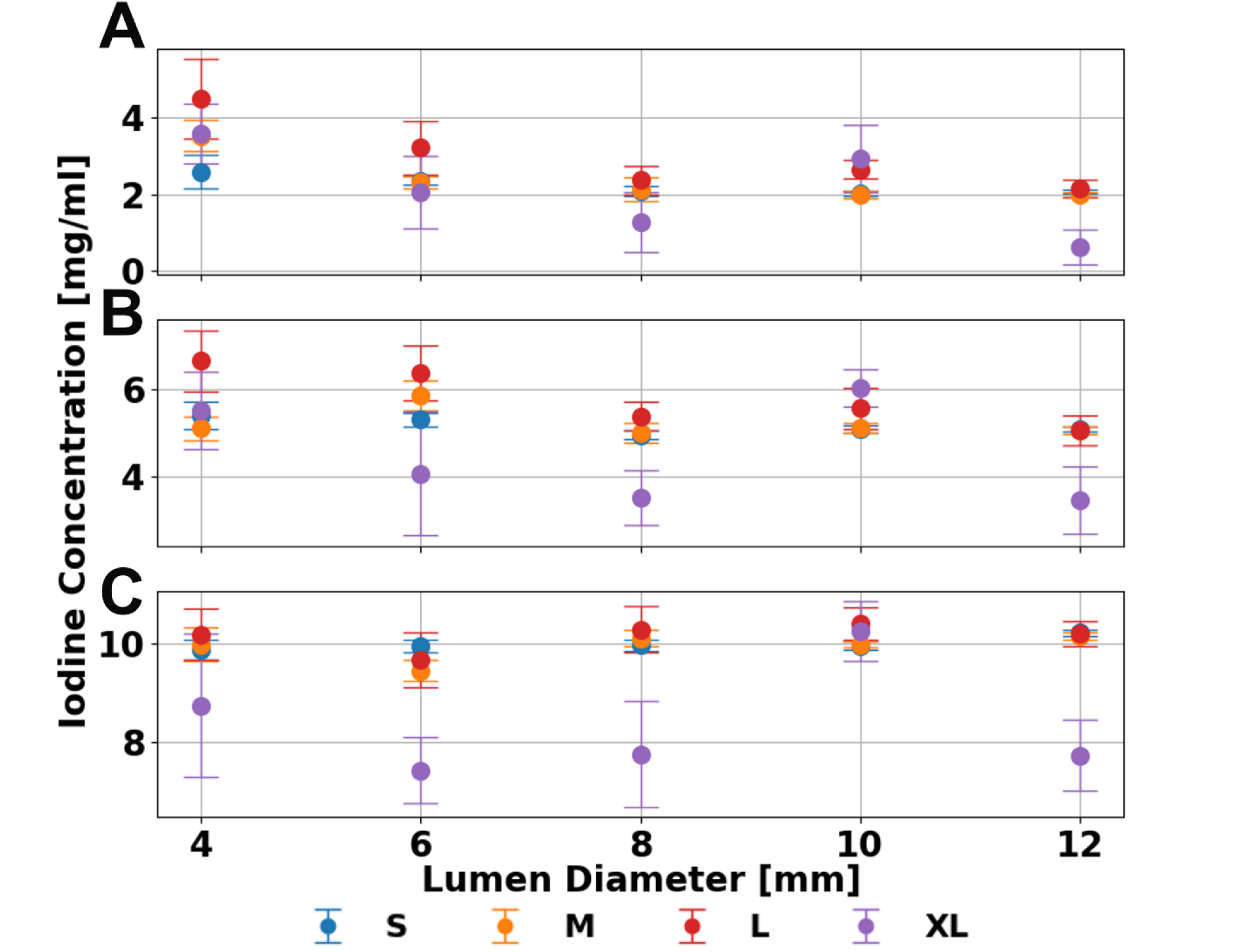
Effect of lumen diameter and phantom size on iodine quantification. Iodine material decomposition remained consistent at most phantom sizes with high accuracy at lumen diameters greater than 6 mm for iodine concentrations of 2 (**A**), 5 (**B**), and 10 (**C**) mg/mL at a radiation dose of 20 mGy. Error bars represent the standard deviation of means collected after ROI placement.

Radiation dose had a limited effect on the accuracy of iodine quantification for lumen diameters larger than 6 mm. Across varying radiation doses, the variance in measured iodine concentration for all phantom sizes and iodine concentrations remained low, with deviations below 0.6 mg/mL. Thus, for larger lumen diameters, iodine quantification remains stable across varying radiation exposure levels, aligning with previous publications^25^.

Radiation dose exerted a more pronounced influence at smaller lumen diameters, specifically for 4 mm inserts, where it dominated over the impact of phantom size. For the 4 mm insert within the L phantom, the absolute relative difference in iodine concentration decreased as radiation dose increased. Specifically, from 5 to 20 mGy, the absolute error in iodine concentration measurements dropped by 1.7 mg/mL, 0.6 mg/mL, and 0.5 mg/mL for iodine concentrations of 2 mg/mL, 5 mg/mL, and 10 mg/mL, respectively. A similar but less pronounced reduction was observed in the S phantom, where absolute error decreased by 0.5 mg/mL, 0.4 mg/mL, and 0.0 mg/mL across the same range of iodine concentrations from 5 to 20 mGy. Moreover, increased radiation dose was associated with reduced measurement uncertainty, as reflected by a decrease in standard deviation (**Figure 4**). For instance, in the L phantom, the average standard deviation across lumen diameters declined by 0.2 mg/mL for each iodine concentration (2 mg/mL, 5 mg/mL, and 10 mg/mL) when radiation dose was increased from 5 to 20 mGy, indicating improved consistency in iodine measurements at higher dose levels.

**Figure 4.**
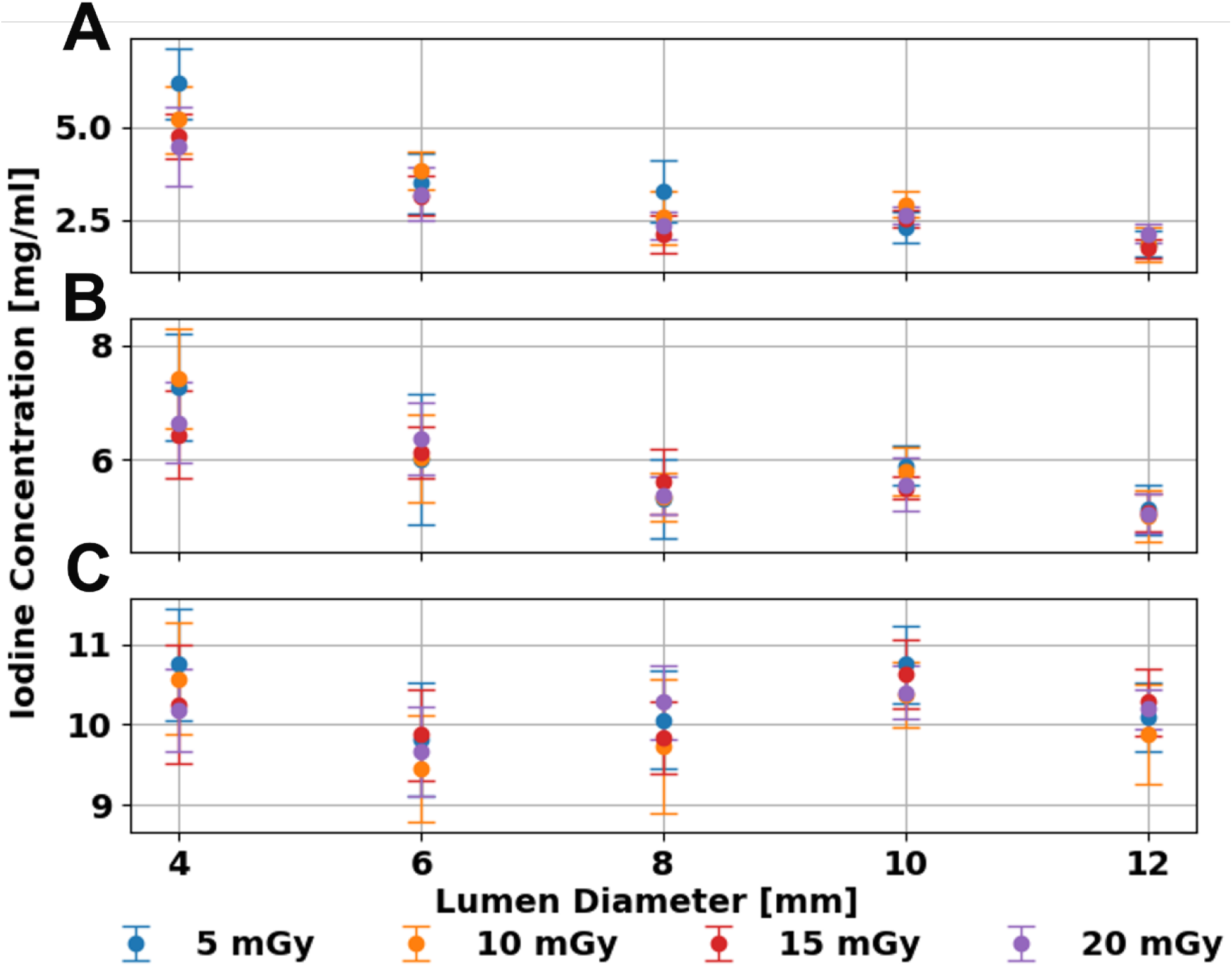
Performance of iodine quantification with a range of radiation doses. Increased radiation dose mitigated the effect of a smaller lumen diameter at phantom size L for iodine concentrations of 2 (**A**), 5 (**B**), and 10 (**C**) mg/mL with error bars representing the standard deviation of ROIs collected at each insert.

The VMI 70 keV images demonstrated stable iodine quantification for lumen diameters above 6 mm, with minimal dependency on iodine concentration or phantom size. Unlike the ID maps, where phantom size influenced measurement accuracy, VMI 70 keV images showed only small changes with phantom size. The average absolute difference across these larger lumen diameters remained below 13 ± 2 HU for all iodine concentrations and phantom sizes. For smaller lumen diameters of 4 mm, however, the dependency on phantom size became more apparent. At a radiation dose of 20 mGy, the absolute difference in VMI 70 keV values increased from 14 HU, 3 HU, and 0 HU for iodine concentrations of 2 mg/mL, 5 mg/mL, and 10 mg/mL in the S phantom to 80 HU, 59 HU, and 38 HU in the XL phantom, respectively (**Figure 5**). Finally, the effect of radiation dose on VMI 70 keV images was minimal for smaller phantom sizes (S and M) and larger lumen diameters (greater than 6 mm), with variances remaining under 3 HU across all iodine concentrations. This stability suggests that, for smaller patient sizes, VMI 70 keV maintains consistent quantification performance without a need for increased radiation exposure.

**Figure 5.**
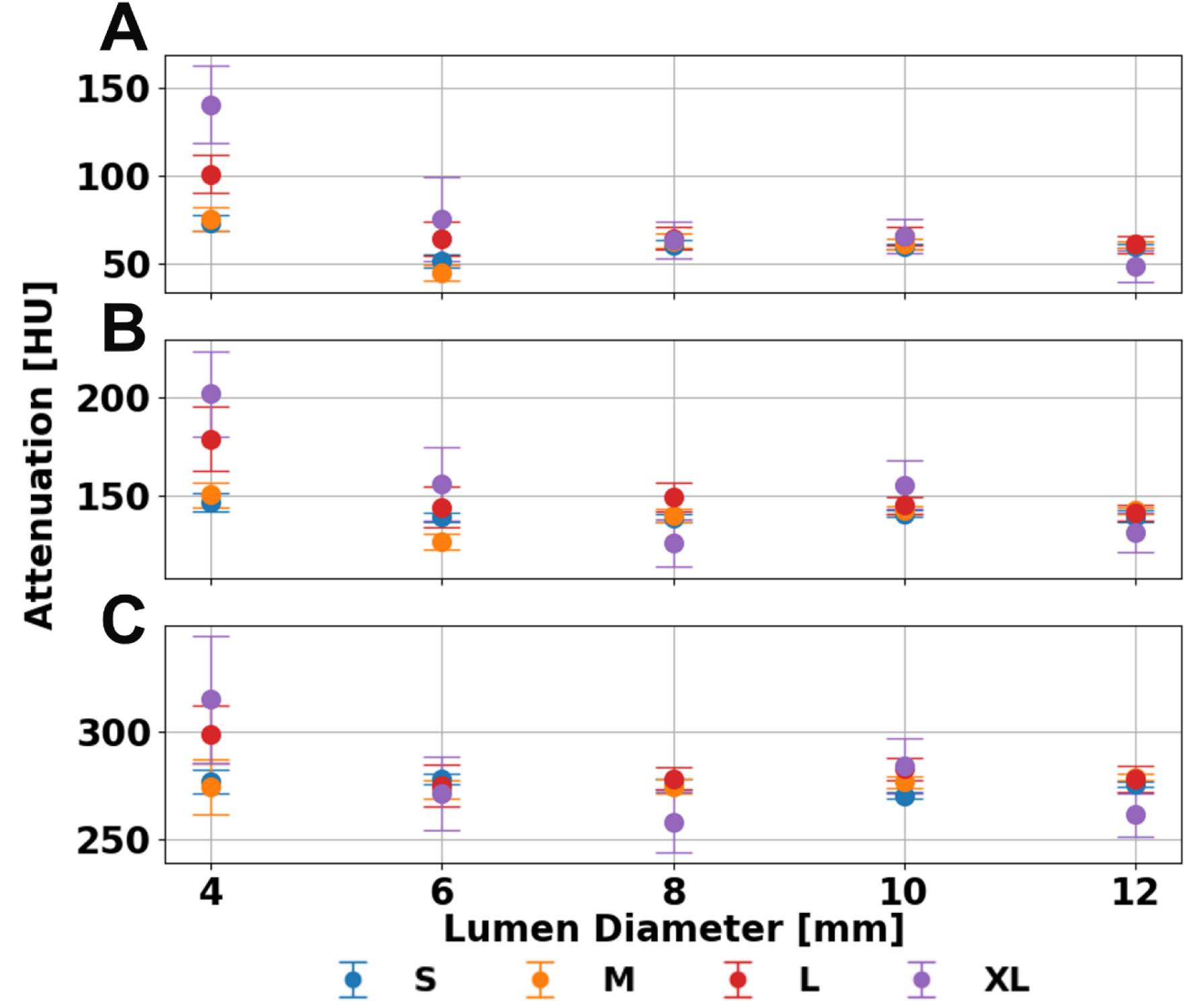
**VMI 70 keV across lumen diameter, phantom size, and iodine concentration**. Error in measured attenuation decreased as lumen diameter increased while phantom size increased the error within the same lumen diameter. Trends with phantom size and lumen diameter were similar for iodine concentrations of 2 (**A**), 5 (**B**), and 10 (**C**) mg/mL. Error bars represent the standard deviation of values measured at each slice.

As with the previous spectral results, VNC images displayed consistent quantification of iodine for lumen diameters greater than 6 mm. Removal of iodine in VNC proved to be accurate for most phantom sizes, except for XL, where mean attenuation showed more error. Average error did not exceed 15 HU for S, M, and L phantom sizes across all iodine concentrations at a dose of 20 mGy. However, the average error at the XL phantom size was 20.2 ± 3.6, 30.2 ± 12, and 51.8 ± 27.4 HU for iodine concentrations of 2, 5, and 10 mg/mL, respectively (**Figure 6)**. The loss of contrast and increased error with increasing phantom size is evident for the 4 mm inserts. As seen in other spectral results, there is an increase in error between the S to XL size where absolute error rises by 60.5 HU, 99.3 HU, and 133.2 HU for concentrations of 2, 5, 10 mg/mL, respectively. This increase in error further illustrates the effect of phantom size on iodine quantification while also demonstrating the viability of iodine removal across most phantom sizes and lumen diameters.

**Figure 6.**
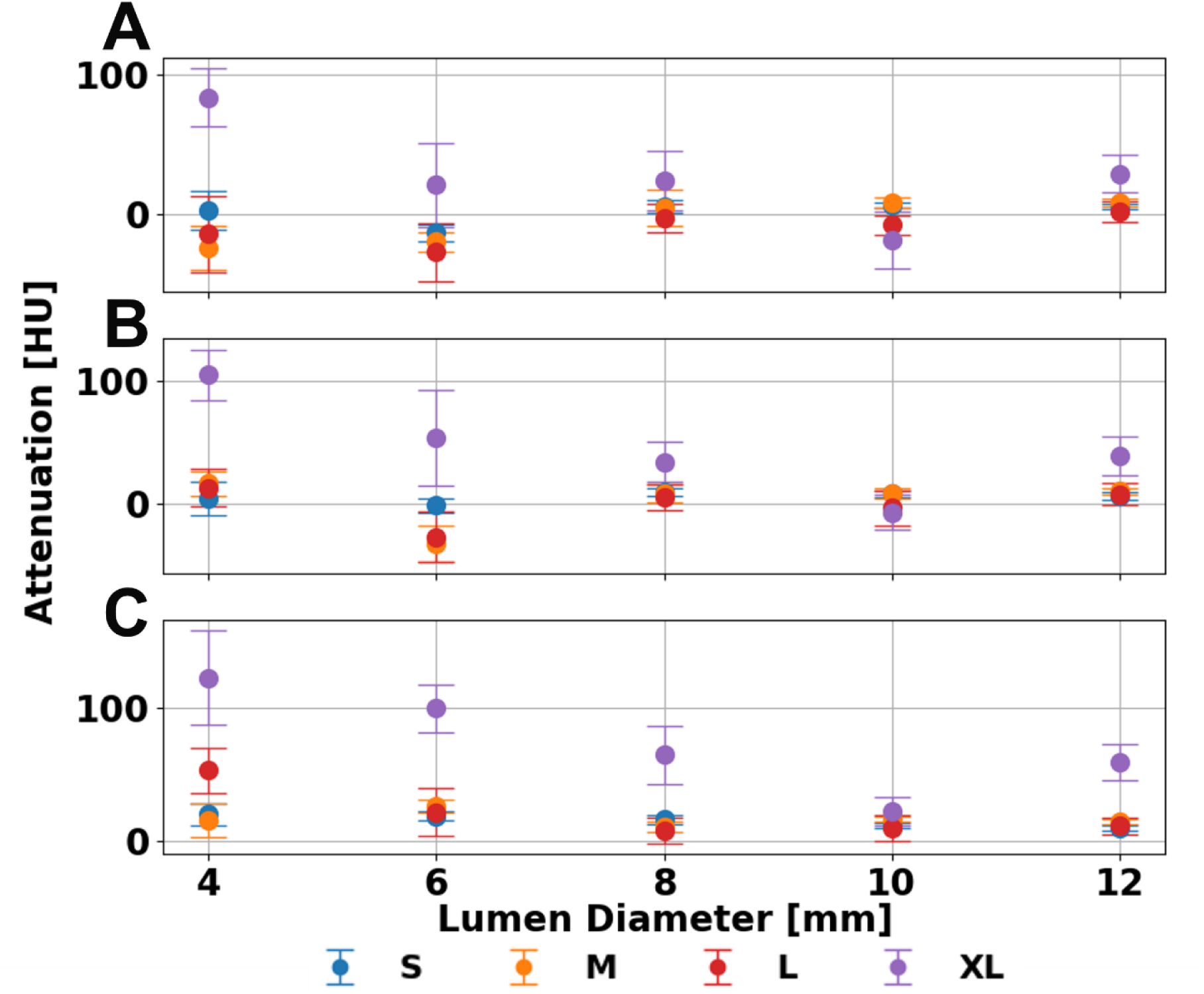
Attenuation after iodine removal across lumen diameter and phantom size. For concentrations of 2 (A), 5 (B), and 10 (C) mg/mL the replacement of iodine to water in VNC images remains consistently accurate for S, M, and L phantom sizes at a CTDIvol of 20 mGy. Error bars represent standard deviation of measured mean attenuation.

## Discussion (670)

Our phantom study provides a comprehensive technical assessment of a clinically available PCCT system, specifically evaluating its ability to accurately quantify iodine in the presence of calcium as well as its overall spectral performance across a diverse range of lumen diameters. The findings from this study offer insights that could inform protocol optimization and guide future research directions for CAD diagnostics and other cardiovascular conditions. Notably, our results highlight that lumen diameter and phantom size influence spectral imaging efficacy. In particular, for lumen sizes exceeding 6 mm, we observed more consistent performance metrics, with both VMI at 70 keV and ID values but exhibiting notable dependencies on phantom size and radiation dose.

Dual-source PCCT holds the potential to enhance cardiovascular imaging by providing superior temporal resolution, reducing motion artifacts, and enabling precise tissue characterization and quantification of materials like iodine and calcium, which are essential for accurate diagnosis and assessment of cardiovascular disease. Our study underscores the importance of lumen size in cardiovascular disease CT diagnostics. Previous studies of PCCT have investigated its potential in diagnosing cardiovascular disease, focusing on the capability to distinguish between calcium and iodine contrast agents through spectral decomposition. For example, van der Werf et al.^19^ examined calcium scoring and demonstrated that PCCT achieves high accuracy in differentiating calcified lesions from adjacent tissues. Furthermore, previous literature^26–28^ has explored the use of VNC images for calcium scoring in PCCT and Dual-energy CT (DECT), recognizing both difficulties—as VNC can underestimate calcification—and its potential to minimize radiation dose exposure by reducing needed scans. Additionally, Dangelmaier et al.^29^ analyzed the separation of iodine, gadolinium, and calcium using PCCT, albeit on an aortic phantom with larger lumen diameters, however, they were able to differentiate calcium and iodine effectively in a similar experimental setting. These findings align closely with our results, which similarly show that PCCT can accurately quantify iodine in this setting.

This study has several limitations that should be taken into account when interpreting the findings. Firstly, the study was conducted using only a single PCCT system, which means that we could not assess inter-scanner variability. Differences between PCCT systems from various manufacturers, could impact performance characteristics and limit the generalizability of our results across other PCCT systems. Secondly, while the PCCT system we used offers an improvement in spatial resolution for spectral data compared to conventional CT, it currently does not simultaneously capture high spatial resolution and spectral data within a single dataset. This limitation restricts the ability to analyze fine anatomical details and spectral information together, which could impact the precision of lumen characterization in clinical practice. Additionally, the phantoms utilized in this study do not fully replicate the anatomical complexity or the diverse composition of plaques found in human patients. Realistic patient anatomy and the variability of plaque composition, which often includes a mixture of calcium, lipid, and fibrous tissue, may introduce challenges not addressed by our simplified phantom model. Moreover, while our study focused primarily on variations in iodine concentration, future research should examine a broader range of calcium concentrations and plaque types to better understand the interplay between calcium and iodine in PCCT imaging. Finally, translating these findings into clinical practice will require further studies involving patient populations. Such clinical trials are essential to confirm the diagnostic value and practical utility of PCCT for assessing cardiovascular disease in real-world settings, where variability in patient anatomy, plaque composition, and clinical presentation may influence imaging performance. Nevertheless, our current study lays the technical groundwork for larger-scale studies.

In conclusion, this study provides a comprehensive technical evaluation of a clinically available PCCT system, emphasizing its ability to generate accurate spectral data for quantifying iodine in the presence of calcium across various lumen diameters and phantom sizes. Notably, the XL phantom, representing the 99th percentile of size of the U.S. population, highlights the system’s capacity to deliver stable and precise imaging across nearly the entire patient size range. These findings offer valuable guidance for optimizing imaging protocols in cardiovascular diagnostics, particularly for CAD, and underscore the potential of PCCT to advance non-invasive cardiovascular imaging.

## Data Availability

All data produced in the present study are available upon reasonable request to the authors.

## Acknowledgments

We acknowledge support from National Institutes of Health (R01EB030494) and Siemens Healthineers.

## References

1. Tsao CW, Aday AW, Almarzooq ZI, et al. Heart Disease and Stroke Statistics—2023 Update: A Report From the American Heart Association. Circulation. 2023;147(8):e93-e621. doi:10.1161/CIR.0000000000001123

2. Weir-McCall JR, Williams MC, Shah ASV, et al. National Trends in Coronary Artery Disease Imaging. JACC Cardiovasc Imaging. 2023;16(5):659–671. doi:10.1016/j.jcmg.2022.10.022

3. Sandfort V, Lima JAC, Bluemke DA. Noninvasive Imaging of Atherosclerotic Plaque Progression: Status of Coronary Computed Tomography Angiography. Circ Cardiovasc Imaging. 2015;8(7):e003316. doi:10.1161/CIRCIMAGING.115.003316

4. Eckert J, Schmidt M, Magedanz A, Voigtländer T, Schmermund A. Coronary CT Angiography in Managing Atherosclerosis. Int J Mol Sci. 2015;16(2):3740–3756. doi:10.3390/ijms16023740

5. Schoenhagen P, Barreto M, Halliburton SS. Quantitative plaque characterization with coronary CT angiography (CTA). Int J Cardiovasc Imaging. 2008;24(3):313–316. doi:10.1007/s10554-007-9284-y

6. McCollough CH, Leng S, Yu L, Fletcher JG. Dual-and Multi-Energy CT: Principles, Technical Approaches, and Clinical Applications. Radiology. 2015;276(3):637–653. doi:10.1148/radiol.2015142631

7. Johnson TRC. Dual-Energy CT: General Principles. Am J Roentgenol. 2012;199(5_supplement):S3-S8. doi:10.2214/AJR.12.9116

8. Greffier J, Villani N, Defez D, Dabli D, Si-Mohamed S. Spectral CT imaging: Technical principles of dual-energy CT and multi-energy photon-counting CT. Diagn Interv Imaging. 2023;104(4):167–177. doi:10.1016/j.diii.2022.11.003

9. Meloni A, Frijia F, Panetta D, et al. Photon-Counting Computed Tomography (PCCT): Technical Background and Cardio-Vascular Applications. Diagnostics. 2023;13(4):645. doi:10.3390/diagnostics13040645

10. Sandfort V, Persson M, Pourmorteza A, Noël PB, Fleischmann D, Willemink MJ. Spectral photon-counting CT in cardiovascular imaging. J Cardiovasc Comput Tomogr. 2021;15(3):218–225. doi:10.1016/j.jcct.2020.12.005

11. Flohr T, Schmidt B, Ulzheimer S, Alkadhi H. Cardiac imaging with photon counting CT. Br J Radiol. 2023;96(1152):20230407. doi:10.1259/bjr.20230407

12. Liu LP, Shapira N, Sahbaee P, et al. Consistency of spectral results in cardiac dual-source photon-counting CT. Sci Rep. 2023;13(1):14895. doi:10.1038/s41598-023-41969-7

13. Si-Mohamed SA, Boccalini S, Lacombe H, et al. Coronary CT Angiography with Photon-counting CT: First-In-Human Results. Radiology. 2022;303(2):303–313. doi:10.1148/radiol.211780

14. van der Bie J, Bos D, Dijkshoorn ML, Booij R, Budde RPJ, van Straten M. Thin slice photon-counting CT coronary angiography compared to conventional CT: Objective image quality and clinical radiation dose assessment. Med Phys. 2024;51(4):2924–2932. doi:10.1002/mp.16992

15. Rajagopal JR, Farhadi F, Richards T, et al. Evaluation of Coronary Plaques and Stents with Conventional and Photon-counting CT: Benefits of High-Resolution Photon-counting CT. Radiol Cardiothorac Imaging. 2021;3(5):e210102. doi:10.1148/ryct.2021210102

16. Holmes TW, Gleason P, Sharma S, et al. Dose-efficient characterization of coronary artery plaques with a prototype CdZnTe-based photon-counting CT scanner. In: Medical Imaging 2024: Physics of Medical Imaging. Vol 12925. SPIE; 2024:557-560. doi:10.1117/12.3008650

17. Symons R, De Bruecker Y, Roosen J, et al. Quarter-millimeter spectral coronary stent imaging with photon-counting CT: Initial experience. J Cardiovasc Comput Tomogr. 2018;12(6):509–515. doi:10.1016/j.jcct.2018.10.008

18. Mannil M, Hickethier T, von Spiczak J, et al. Photon-Counting CT: High-Resolution Imaging of Coronary Stents. Invest Radiol. 2018;53(3):143–149. doi:10.1097/RLI.0000000000000420

19. van der Werf NR, Booij R, Greuter MJW, et al. Reproducibility of coronary artery calcium quantification on dual-source CT and dual-source photon-counting CT: a dynamic phantom study. Int J Cardiovasc Imaging. 2022;38(7):1613–1619. doi:10.1007/s10554-022-02540-z

20. Eberhard M, Mergen V, Higashigaito K, et al. Coronary Calcium Scoring with First Generation Dual-Source Photon-Counting CT-First Evidence from Phantom and In-Vivo Scans. Diagn Basel Switz. 2021;11(9):1708. doi:10.3390/diagnostics11091708

21. Symons R, Sandfort V, Mallek M, Ulzheimer S, Pourmorteza A. Coronary artery calcium scoring with photon-counting CT: first in vivo human experience. Int J Cardiovasc Imaging. 2019;35(4):733–739. doi:10.1007/s10554-018-1499-6

22. van der Werf NR, Si-Mohamed S, Rodesch PA, et al. Coronary calcium scoring potential of large field-of-view spectral photon-counting CT: a phantom study. Eur Radiol. 2022;32(1):152–162. doi:10.1007/s00330-021-08152-w

23. Rajendran K, Petersilka M, Henning A, et al. First Clinical Photon-counting Detector CT System: Technical Evaluation. Radiology. 2022;303(1):130–138. doi:10.1148/radiol.212579

24. Liu LP, Shapira N, Chen AA, et al. First-generation clinical dual-source photon-counting CT: ultra-low-dose quantitative spectral imaging. Eur Radiol. 2022;32(12):8579–8587. doi:10.1007/s00330-022-08933-x

25. Liu LP, Pua R, Dieckmeyer M, et al. Impact of patient habitus and acquisition protocol on iodine quantification in dual-source photon-counting computed tomography. J Med Imaging Bellingham Wash. 2024;11(Suppl 1):S12806. doi:10.1117/1.JMI.11.S1.S12806

26. Dobrolinska MM, Koetzier LR, Greuter MJW, et al. Feasibility of virtual non-iodine coronary calcium scoring on dual source photon-counting coronary CT angiography: a dynamic phantom study. Eur Radiol. 2024;34(11):7429–7437. doi:10.1007/s00330-024-10806-4

27. Sharma SP, van der Bie J, van Straten M, et al. Coronary calcium scoring on virtual non-contrast and virtual non-iodine reconstructions compared to true non-contrast images using photon-counting computed tomography. Eur Radiol. 2024;34(6):3699–3707. doi:10.1007/s00330-023-10402-y

28. Gassert FG, Schacky CE, Müller-Leisse C, et al. Calcium scoring using virtual non-contrast images from a dual-layer spectral detector CT: comparison to true non-contrast data and evaluation of proportionality factor in a large patient collective. Eur Radiol. 2021;31(8):6193–6199. doi:10.1007/s00330-020-07677-w

29. Dangelmaier J, Bar-Ness D, Daerr H, et al. Experimental feasibility of spectral photon-counting computed tomography with two contrast agents for the detection of endoleaks following endovascular aortic repair. Eur Radiol. 2018;28(8):3318–3325. doi:10.1007/s00330-017-5252-7

